# Point of care testing for detection of coronaviruses including SARS-CoV-2 from saliva without treating RNA in advance

**DOI:** 10.1101/2021.05.01.21256441

**Authors:** Masaaki Muraoka, Osamu Kawaguchi, Mikio Mizukoshi, Shunsuke Sejima

## Abstract

Coronavirus disease 2019 (COVID-19) outbreak was reported to the WHO (World Health Organization) as an outbreak on end of 2019, afterwards pandemic on the worldwide in 2020. Severe acute respiratory syndrome coronavirus 2 (SARS-CoV-2) has be reported it to cause COVID-19, and highly transmissible. Therefore, it is important that it is rapidly and continuously detected and monitored on site so that the infection is prevented. Namely, POCT (point of care testing) may be important to control cross infection of SARS-CoV-2. At present, the routine confirmation of SARS-CoV-2 is based on detection of sequence unique in the virus RNA by nucleic acid amplification tests (NAATs) such as rRT-PCR. Taking POCT into account, it is clear that it takes time and labour very much or much. Thus, it was our purpose this time to contribute to develop POCT for microbes such as SARS-CoV-2, and thus needed to improve NAATs method.

First, combining the mobile real-time RT-PCR (rRT-PCR) device PCR1100 with the appropriate rRT-PCR reagent, we found that it is possible to detect RNA of SARS-CoV-2 for less 14 minutes with equivalent accuracy to conventional devices. Next, we found that the above method made it possible for us to detect coronaviruses by direct rRT-PCR without pre-treatments. Furthermore, it also made clear that coronaviruses in saliva could be detected by the similar direct rRT-PCR method.

Hence, it was concluded that this method made it possible to detect virus in saliva without treating in advance (extraction, purification, concentration, etc.), and moreover, samples would be able to be collected with non-invasive. For this reason, we suggest that this method is useful for POCT of coronaviruses including SARS-CoV-2.

## INTRODUCTION

Severe acute respiratory syndrome-coronavirus 2 (SARS-CoV-2) is a cause of coronavirus disease 2019 (COVID-19) outbreak, of which the WHO (World Health Organization) declared pandemic on March 11, 2020 [1]. COVID-19 was firstly reported to the WHO as an outbreak in Wuhan City, Hubei Province, China on December 31, 2019, afterwards has rapidly spreading [2]. Human infectious coronaviruses, including SARS-CoV-2, have been confirmed seven types as of this moment. Four out of them are causes of common cold, and as follows: human coronavirus (HCoV)-229E, -HKU1, -OC43, and -NL63. Meanwhile, other three human coronaviruses (middle east respiratory syndrome coronavirus [MERS-CoV], SARS-CoV, SARS-CoV-2) are highly pathogenic, and can cause severe diseases presented as acute respiratory distress syndrome (ARDS) characterized by dysfunctional immune responses and severe pulmonary injury [3, 4]. SARS-CoV-2 is genetically closely related to SARS-CoV, however which is less deadly and more transmissible than MERS-CoV and SARS-CoV [5]. Therefore, it is important that it is rapidly and continuously detected and monitored on site so that the infection is prevented. Namely, point of care testing (POCT) may be important to control cross infection of SARS-CoV-2 as other studies reported [6, 7].

At present, the routine confirmation of SARS-CoV-2 is based on detection of sequence unique in the virus RNA by nucleic acid amplification tests (NAATs) such as rRT-PCR [8], for various target sites of which have reported to the following: nucleocapsid (N), envelop (E), RNA-dependent RNA polymerase (RdRp)/open reading frame 1 ab (Orf1ab), and spike (S) [9, 10, 11, 12, 13]. As described above, we recognize that the performance characteristics of NAATs can vary with reagents, programs, and devices, and moreover, that conventionally, the rRT-PCR requires more than 60 minutes for RNA extraction, etc., and further 90 minutes for transcription and amplification [14, 15, 16]. Thus, it is clear that it must take much time and much labour. Incidentally, Nasopharyngeal and oropharyngeal swabs are the recommended specimen types for SARS-CoV-2 testing by NAATs [17, 18]. However, these sampling methods pose a risk of cross infection between patients and healthcare workers, since these cause discomfort such as coughing and sneezing. Furthermore, these sampling may also cause bleeding, so that is invasive for patients. By contrast, saliva collection causes less patients discomfort, and moreover, less possible cross infection because it may be useful for self-collection [19, 20, 21].

It was our purpose this time to contribute to develop POCT for microbes such as SARS-CoV-2, and thus needed to improve NAATs method. Therefore, it was first focused on the mobile rRT-PCR device ’PCR1100’
s and rRT-PCR reagent kit and evaluated whether they were ’rapid and easy’, similarly to recently other reports for detection of SARS-CoV-2 [12] and dengue virus [22]. Next, it was considered whether rRT-PCR could be performed in saliva specimen or not without treating RNA extraction, etc. in advance so-called ’direct’. HCoV-229E was substituted for SARS-CoV-2 to evaluate this. Because HCoV-229E is easily accessible for the biosafety level 2 (BSL2) laboratory, HCoV-229E may be a good initial model for the evaluation against other coronaviruses, such as SARS-COV-2 with high-risk for biosafety [23].

## MATERIALS AND METHODS

### ETHICS STATEMENT

The study was approved by the Ethics Committee of Certified Non-Profit Organization Biomedical Science Association, Japan (BMSA2020-12-01) for the evaluation of detecting HCoV-229E in saliva. Spoken informed consent was obtained from each volunteer participated to offer saliva.

### Sample

Positive control RNA of SARS-CoV-2 for the NIID protocol were kindly provided from NIID (for N primer probe Ver.2 and N2 primer probe Ver.2/Ver.3). For the CDC protocol were synthesised (Fasmac Co. Ltd., Japan) based on each primer sequence and information of GenBank™: accession number MN997409.1, position −10/+110 for 2019-nCoV_N1 and position +871/+980 for _N2. Each positive control RNA was applied 10-fold serial dilution with 10mM Tris-HCl (pH8.0, molecular grade) included 10 µg/mL carrier RNA (Ribonucleic acid from baker’s yeast, Merck KGaA, Germany).

HCoV-229E was purchased (VR-740™, American Type Culture Collection [ATCC]^®^, USA), followed by infection with MRC-5 cells (CCL-171™, ATCC^®^) and proliferating with EMEM medium (30-2003™, ATCC^®^) included 2 % FBS (30-2020™, ATCC^®^) for 5 to 8 days in 5 % CO_2_ at 35 °C. Only the supernatant was utilized in each assay without treating in advance, that was, evaluated for rRT-PCR with keeping it raw. Virus titers (median tissue culture infectious dose [TCID50/mL]) was determined by the conventional method in each evaluation. Subsequently, to evaluate the correlation of detection value on the device PCR1100 with virus titers, the non-treated sample was applied 30-fold serial dilution with 10mM Tris-HCl (including 10 µg/mL carrier RNA).

Saliva samples were offered from volunteers. Volunteers consisted of forties regardless of gender and whether infected by human coronavirus. Each saliva sample was collected by rinsing intraoral with 2mL of saline (API^®^ NaCl 0.85% Medium, bioMérieux SA, France) for 15 seconds or more, and expelled to the appropriate container following by utilizing to dilute HCoV-229E.

### Primer and Probe

Based on previously reported protocols, for SARS-CoV-2 by the NIID protocol [10, 11] and the CDC protocol [9, 13], and moreover, for HCoV-229E [15], respectively, oligonucleotide primers and probes for rRT-PCR detection were selected to target each N gene and synthesised (Nihon Gene Research Laboratories Inc., Japan) (Table 1). The fluorescent dyes attached with the 5’ end of each probe were the following: for N_Sarbeco_P, with 6-carboxyfluorescein (FAM); for NIID_2019-nCOV_N_P2, with cyan5 (Cy5); for 2019-nCoV_N1-P, with FAM; for 2019-nCoV_N2-P, with 6-carboxy-2’
s,4,4’,5’,7,7’-hexachlorofluorescein (HEX); and for HCoV-229E_NP, with FAM. ALL probes were attached with Black Hole Quencher (BHQ, Biosearch Technologies, Inc., USA) at the 3’ end.

**Table 1.**
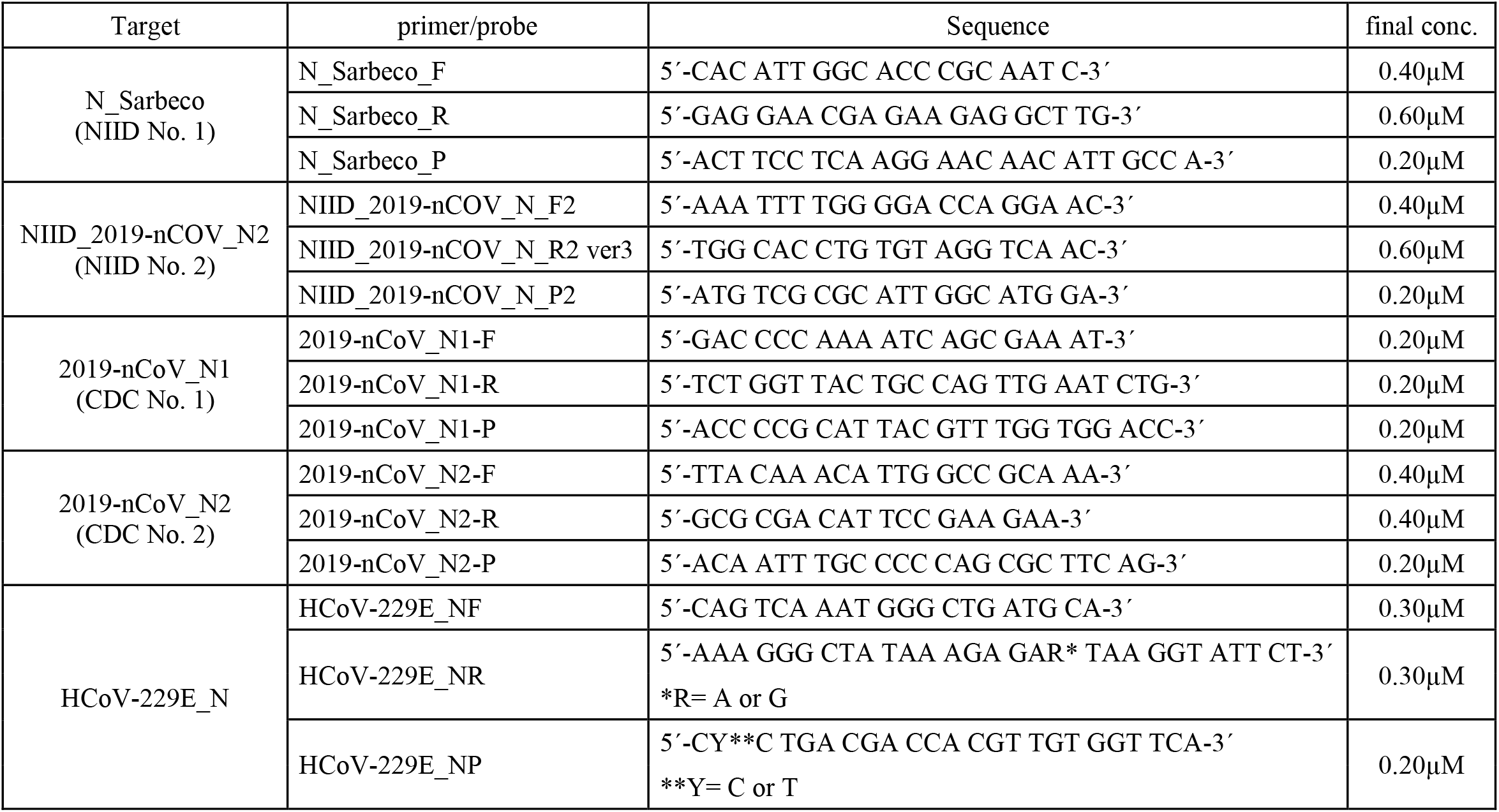
The list of oligonucleotide primer and probe. final conc.: final concentration in RT-PCR.

### Real-time RT-PCR

RT and PCR were performed in the one-step with utilizing the device PCR1100 for all tests of this study. Briefly describe the composition of rRT-PCR reagent, 3 µL of a sample (the raw sample or positive control RNA) was amplified in a 17 µL reaction solution containing 1x Reaction Buffer, 0.25 µL or 0.50 µL of RT Enzyme Mix, 1.0 µL of DNA Polymerase (THUNDERBIRD^®^ Probe One-step qRT-PCR Kit, TOYOBO Co. Ltd., Japan), each final concentration of primer and probe for target detection (Table 1). With preliminary tested, each composition optimized. As a result, the adequate volume of RT Enzyme Mix was 0.50 µL in all protocol other than CDC protocol.

Each rRT-PCR condition adequate for the device PCR1100 in this study was programmed as follows: RT incubation was followed by enzyme activation, so which was performed first at 50 °C for 150 seconds, next at 95 °C for 15 seconds. Afterwards, denaturation, and annealing and amplification was performed for fifty cycles as follows: for SARS-CoV-2 based by the NIID protocol, at 95 °C for 3.5 seconds, and at 60 °C for 16 seconds; for SARS-CoV-2 based by the CDC protocol, at 95 °C for 3.5 seconds, and at 60 °C for 7 seconds; and for HCoV-229E, at 95 °C for 3.5 seconds, and at 60 °C for 8 seconds. With preliminary tested, each condition was optimized. Incidentally, the procedure of the device PCR1100 was according to previously reported [24].

## RESULTS

### Detection of SARS-CoV-2 in positive control RNA

To begin with, in the NIID protocol, the positive control RNA could be simultaneously detected by both NIID No. 1 and NIID No. 2 primer/probe set with multichannel (Fig. 1). Moreover, each Ct value was correlatively depended on the rate of serial dilution (R^2^ > 0.99). Meanwhile, also in the CDC protocol, it was possible for us to detect with multichannel of CDC No. 1 and CDC No. 2 simultaneously, and moreover, each Ct value was correlatively shifted to the rate of serial dilution (R^2^ > 0.99).

**Fig. 1.**
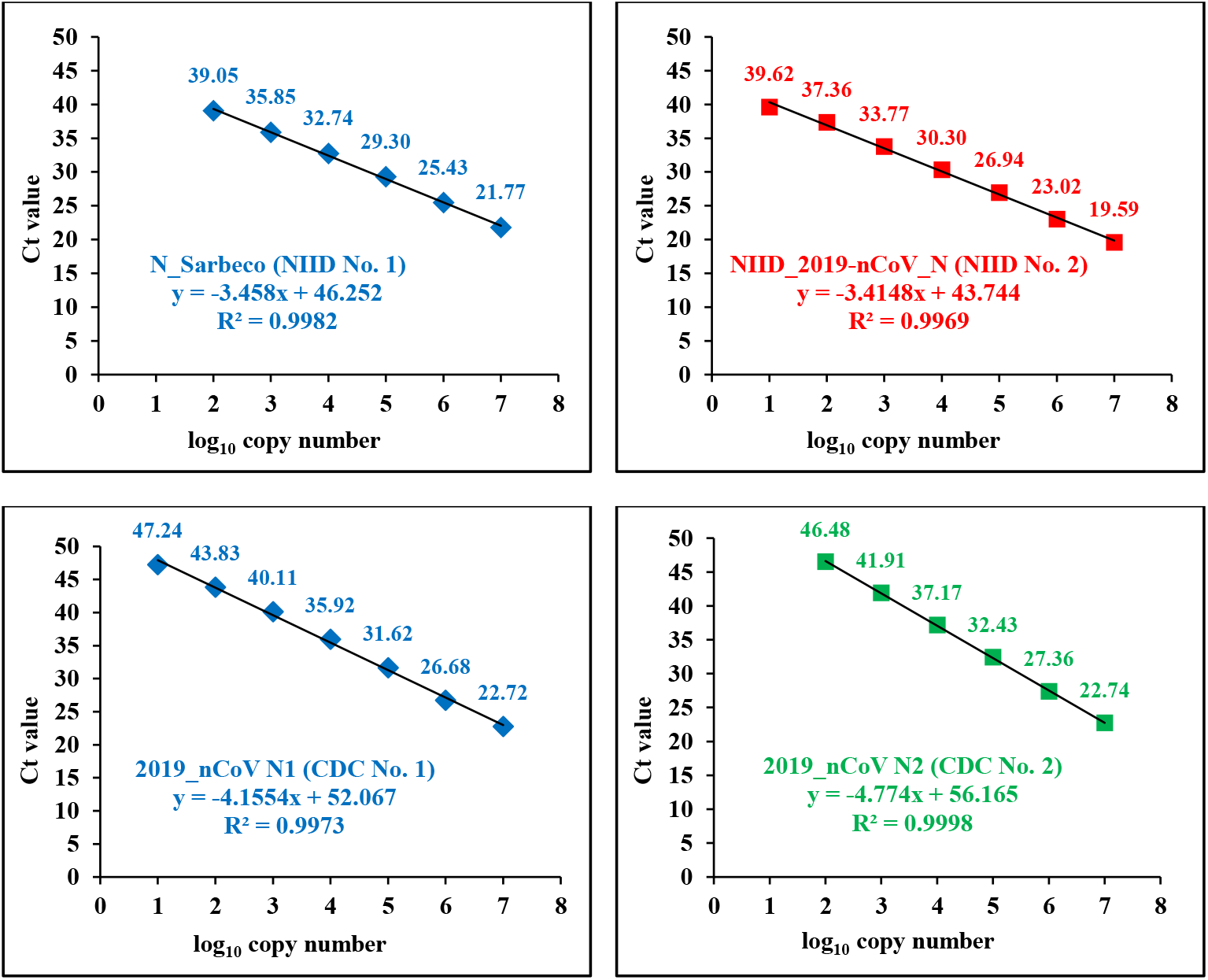
Correlation of RNA copy number and Ct value when multichannel detection of positive control: The upper left and right figure in the NIID protocol, and the lower left and right figure in the CDC protocol show high correlation of copies number to Ct value, respectively. x: log_10_ copy number (per reaction), y: Ct value, numeral on each marker: Ct value against each copy number.

In the limit of detection (LOD) of the NIID protocol, the NIID No. 2 was detectable more sensitive than NIID No. 1, and 10 copies/reaction could be determined. Meanwhile, in the LOD of the CDC protocol, the CDC No. 1 was detectable more sensitive than CDC No. 2, and 10 copies/reaction could be determined.

Next, when the NIID protocol compared to the CDC protocol in detection time, the NIID protocol took time more than the CDC protocol the following: NIID, for 20 minutes; and CDC, for 13.5 minutes. For this reason, it was found that when CDC protocol No. 1 and No. 2 primer/probe set was utilized by this method simultaneously, measurement values could be provided with rapid and accurate method.

### Detection of HCoV-229E without treating in advance

First, to evaluate whether rRT-PCR could be performed by the above method without treating in advance, HCoV-229E was utilized instead of SARS-CoV-2. Incidentally, the virus titer of utilized HCoV-229E was 7.2 x 10^6 TCID50/mL this time which was diluted and performed to rRT-PCR with non-treated. As a result, 30-fold serial dilution with 10mM Tris buffer was highly correlated to Ct value (Fig. 2, R^2^ > 0.99). As a result, without treatment for extraction, concentration and purification for RNA, it was possible for us to detect HCoV-229E. Besides, the LOD was at least 8.9 x 10^2^ TCID_50_/mL, and it took only approximately 13 minutes per detection by utilizing the device PCR1100 with one step rRT-PCR reagent the same above of SARS-CoV-2.

**Fig. 2.**
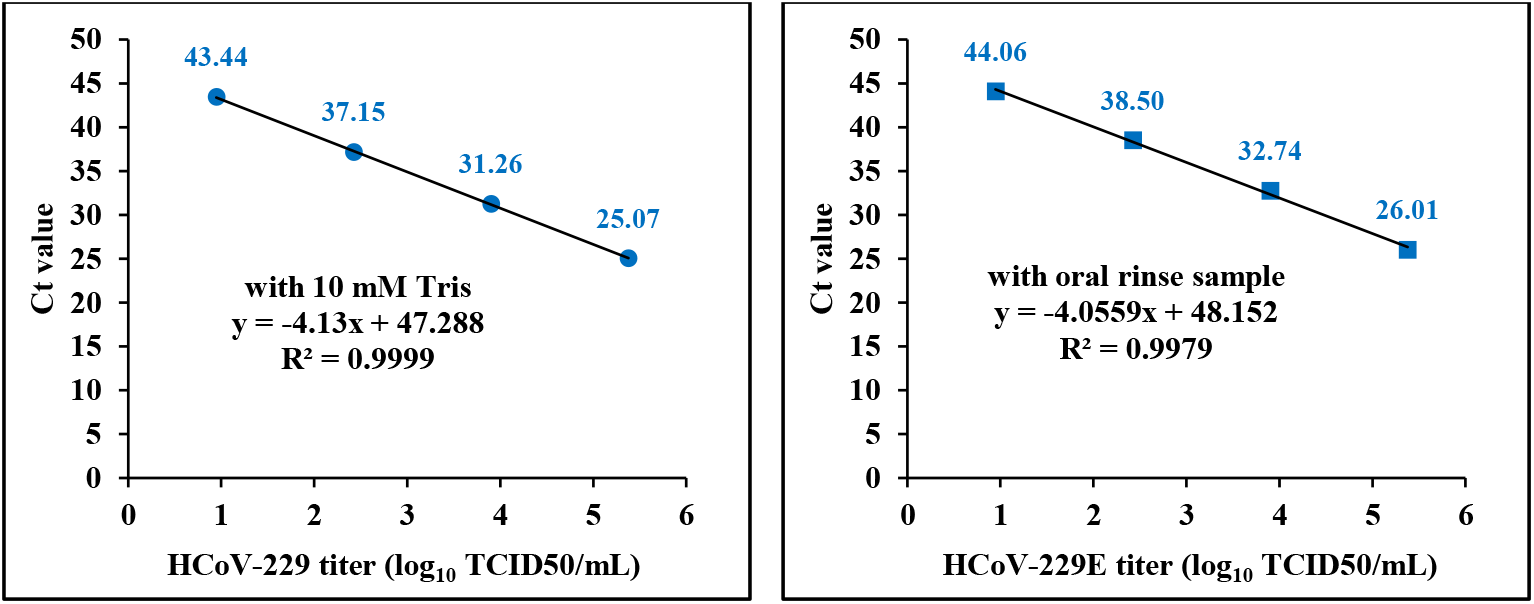
Correlation of virus titer and Ct value when detection of HCoV-229E by real-time direct RT-PCR: The left figure shows result of serial dilution with 10mM Tris-HCl, and the right figure show result of serial dilution with saliva sample (expelled from the mouth after rinsing), respectively, with high correlation of virus titer to Ct value. x: log_10_ TCID50/mL, y: Ct value, numeral on each marker: Ct value against each titer.

Next, we further assessed whether it was possible to detect virus in saliva without treating in advance. Accordingly, the saliva sample (expelled from the mouth after rinsing) instead of 10 mM Tris buffer was utilized to dilute HCoV-229E followed by performing to rRT-PCR without treating in advance. As a result, it was identified that HCoV-229E could be detected in saliva sample with non-treating, and moreover, 30-fold serial dilution was highly correlated to Ct value (Fig. 2, R^2^ > 0.99), similarly to 10 mM Tris buffer.

## DISCUSSION

As several studies have reported [12, 24, 25], with utilizing the mobile real-time PCR device PCR1100, it is possible to detect organism rapidly and easily compared to other conventional devices. Conceiving POCT, however, there still left so much time and much labour things to improve to detect SARS-CoV-2. Therefore, we developed and evaluated more rapid and easier method.

First, our aim was to establish the new protocol to detect SARS-CoV-2 rapidly; therefore, the reagent was selected to be able take advantage of characteristics of the rRT-PCR device PCR1100. The selected rRT-PCR reagent makes it possible to perform RT and PCR by one-step continuously, so that it omits extra labor and time [26]. Furthermore, about primer and probe, the time required for PCR was conceived to shorten. Consequently, it was focused on the CDC protocol, and compared to the NIID protocol since both protocols target two sets for N-gene region [9, 11]. As a result, it was clear that both of the CDC protocol and the NIID protocol made it possible for us to detect RNA of SARS-CoV-2 with the each multichannel, simultaneously No.1 and No. 2 (Fig. 1). Moreover, although the CDC protocol was equivalent LOD to the NIID protocol (from 10 to 100 copies), it took less 14 minutes for measurement in the CDC protocol in contrast to the NIID protocol which it took 20 minutes. The present work demonstrated that the NIID protocol made it for each Ct value to correlate to the serial dilution highly, similarly to other studies [11]. On the other hand, it was demonstrated results of the CDC protocol was also similar to the NIID protocol. In the context of these reasons, it was suggested that when the CDC protocol was utilized by this multichannel method, SARS-CoV-2 could be detected with more rapidity and equal accuracy as compared to conventional methods.

Next, the most important point that required clarification in this work was that direct rRT-PCR could be performed to the virus in the intended sample. Therefore, by the above method based on the combination of the rRT-PCR device PCR1100 with the rRT-PCR reagent, it was evaluated whether direct rRT-PCR could be performed for HCoV-229E instead of SARS-CoV-2. As a result, it was found that the above method made it possible for us to detect HCoV-229E with direct rRT-PCR (Fig 2). However, in the medium sample, it was unclear which it was possible to detect virus or RNA without treating in advance.

Meanwhile, in line with other studies [19, 20, 21] that it was possible to detect coronavirus in saliva sample rapidly and easily, we focused on intraoral sample, saliva; therefore, the saliva sample (expelled from the mouth after rinsing) was utilized to dilute HCoV-229E followed by performing to direct rRT-PCR, and it was evaluated. As a result, serial dilution rate with the saliva sample was highly correlated to Ct value with equivalent to Tris buffer. As several studies have reported, salivary RNA is prone to degradation by RNases and other nucleases present in saliva [27, 28]. Hence, it was concluded that this method made it possible to detect virus in saliva without treating in advance (extraction, purification, concentration, etc.), and moreover, samples would be able to be collected with non-invasive. We performed rinsing intraoral with 2 mL of saline for 15 seconds to collect saliva this time, however, further consideration will be needed to yield appropriate conditions about POCT. Furthermore, other studies reported that viral load of SARS-CoV-2 in saliva was different with the phase after infection and the symptom [29, 30]; therefore, the detection of SARS-CoV-2 from saliva or others in patients would be required for confirming this in future.

## Data Availability

Data openly available in a public repository that issues datasets with DOIs

## STUDY DESIGN

When each sample was assessed in the real-time RT-PCR, dilution series of each was indicated with high relativity to Ct value on the mobile real-time PCR device PCR1100 (R^2^ > 0.99).

## AUTHOR CONTRIBUTIONS

M. Muraoka conceived, designed and performed experiments, analysed the data and wrote the manuscript. O. Kawaguchi conceived and designed experiments. M. Mizukoshi conceived and provided specimens. All authors also participated in the editorial process and approved the manuscript.

## ACKNOWLEDGEMENTS

We thank volunteers participated to offer saliva, Dr. Kazuya Shirato and Dr. Tsutomu Kageyama at NIID for positive control, and Takashi Fukuzawa in an ex-employee at Nippon Sheet Glass Co. Ltd. for technical comments. Furthermore, this journal is based by own preprint, which we have registered with medRxiv [31], so that we thank medRxiv for the opportunity to submit as the preprint.

